# Utilization Analysis and Fraud Detection in Medicare via Machine Learning

**DOI:** 10.1101/2024.12.30.24319784

**Authors:** Mahan Tajrobehkar, Xin Guo, Diane Nguyen, Nikita Chigullapally, Varun Shravah, Shannon Yee, Abishek Chozhan, Kathy Hoang, Robert T. Chang, Carolina Gutierrez, Scott Lee

## Abstract

Healthcare fraud and overutilization pose significant challenges in the United States, leading to substantial financial losses and compromised patient care. Medicare, a vital federal healthcare program, is particularly susceptible to such abuses. With over 63 million Americans enrolled in Medicare and growing expenses, the need for effective fraud detection is paramount. Traditional methods relying on manual audits have proven insufficient, allowing a significant portion of fraudulent activity to go undetected. Machine learning (ML), however, has gained significant attention in recent years due to its potential for improving the efficiency of fraud detection and prevention. Nonetheless, there are several issues with the existing studies utilizing ML that limit their effectiveness. The most common issue is the heavy reliance on the List of Excluded Individuals and Entities (LEIE) from the Office of the Inspector General for model training and evaluation. Apart from the severe class imbalance issue (with a fraud rate between 0.038% and 0.074%), another notable problem associated with using the LEIE dataset is that many of the providers listed there were prosecuted due to overt and deliberate fraudulent billing. Consequently, using this dataset to train ML models can help detect brazen, outlandish billing patterns, but would be unable to pinpoint instances of more subtle fraud from which a majority of the financial loss and waste occurs. In this paper, we leverage the experience of seasoned physicians and medical billers to create a labeled dataset that overcomes the issues of class imbalance and the exclusive focus on overtly fraudulent providers. We leverage our access to domain knowledge by focusing on the field of ophthalmology. Additionally, using the labeled dataset, we conduct a comparative study of various machine learning models for the task of predicting Medicare overutilization within ophthalmology. The results indicate that our proposed ensemble outperforms individual models such as extreme gradient boosting and multilayer perceptron in detecting overutilization, achieving Area Under the Receiver Operating Characteristic Curve (AUROC score) of 0.907. By deploying the stacking ensemble model, our paper estimates nationwide and jurisdiction-specific overutilization rates, revealing that approximately 8.6% of ophthalmologists engaged in overutilization practices in 2021. We also highlight potential monetary losses of $437.1 million attributed to overutilization activities within ophthalmology for that year alone. Furthermore, feature importance analysis using SHAP (SHapley Additive exPlanations) values provides insights into the key factors influencing the model’s overutilization predictions. Notably, the ratio of total Medicare payments to the total number of patients emerges as a crucial feature in identifying potential overutilizers.

## 1 Introduction

It is estimated that over $100 billion is lost each year in the United States due to healthcare fraud and abuse accounting for 3-10% of all healthcare expenditures [30, 5]. One primary target for overutilization and fraud is Medicare, for which utilization data is publicly available. Over 63 million Americans are currently enrolled in Medicare and the total cost of the program in 2021 is $900.8 billion, accounting for 21% of total national healthcare expenditures[36]. Given a rapidly aging American population, 75 million Americans will be enrolled in Medicare by 2027, and Medicare spending is projected to jump to nearly $1.6 trillion by 2028[16, 22]. Despite the increase in federal healthcare spending over past decades, patients are being forced to shoulder increasingly more of the financial burden. Between 2002 and 2022, Medicare Part B deductibles have risen 133% and monthly premiums have risen 215%[32, 35]. Yet, even with increased spending from both the federal government and patients, the Medicare trust fund is projected to become insolvent in the near future.[25]

A small fraction of medical providers are known to have engaged in various forms of overutilization or fraud to exploit the Medicare system. One prevalent form is phantom billing, where unnecessary procedures are performed and billed to Medicare. Another form is upcoding, where providers bill for higher reimbursing procedures while actually performing lower-reimbursing ones. Additionally, cloned documentation occurs when providers copy information from previous patient records or even from different patients, leading to false or inaccurate billing. These practices not only waste healthcare resources but also compromise patient care, and even put lives at risk with unnecessary procedures. Currently, there is no reliable method to detect overutilization or fraud in healthcare. Fraud detection within healthcare is primarily done through a manual effort by auditors and investigators searching through billing records in a time-consuming and often imprecise manner. It often depends on whistleblowers or insiders to report employers, and therefore the vast majority of overutilization and fraud goes undetected. In 2019, only $2.6 billion of fraudulent claims were recovered by the United States Department of Justice [34]. Given the lack of domain expertise and the subtlety of Medicare abuse by healthcare providers, it can be difficult for auditors to determine what test or procedure is medically necessary in a particular office population.

Detection of fraud using machine learning (ML) has gained significant attention in recent years due to its potential for improving the efficiency and effectiveness of fraud identification and prevention. Since the Centers for Medicare and Medicaid Services (CMS) released public data files in 2014, numerous studies have explored the application of various ML techniques in Medicare fraud detection. For instance, Bauder and Khoshgoftaar [10] conducted a comparative study with supervised, unsupervised, and hybrid ML approaches. They found that the supervised methods tend to perform better than unsupervised or hybrid methods, but the results could vary depending on the class imbalance sampling technique and provider type. Herland et al. [15] focused on the detection of Medicare fraud using various CMS public datasets. They provided detailed discussions on Medicare data processing and exploratory analyses in order to show the best learners and datasets for the detection of fraudulent claims. Several studies had a narrower focus on the performance and applications of supervised ML algorithms such as gradient boosted decision trees (Hancock and Khoshgoftaar [26]), CatBoost (Hancock and Khoshgoftaar [21]), bagging (Yao et al. [27]), and deep learning (Johnson and Khoshgoftaar [19]; Mayaki and Riveill [29]) in identifying Medicare fraud. Additionally, there have been works that deployed unsupervised approaches to identify fraudulent outliers and Medicare anomaly. (Bauder and Khoshgoftaar [6]; Branting et al. [7]; Bauder et al. [13]; Sadiq and Shyu [20])

While ML techniques have shown promise in detecting Medicare fraud, there are several issues that can limit their effectiveness. A common issue with existing studies is the heavy reliance on the List of Excluded Individuals and Entities (LEIE) [33] for model training and evaluation. LEIE is a database compiled by the Office of the Inspector General that reports individuals and entities that have been excluded from receiving federally-funded healthcare programs due to fraud. A significant hurdle in using known fraud labels from the LEIE dataset is the issue of class imbalance with a fraud rate between 0.038% and 0.074%. Data-level techniques such as random sampling and varying class distribution are shown to help mitigate the issue of class imbalance (Brauder and Khoshgoftar [12, 14]; Johnson and Khoshgoftaar [19]; Hancock et al. [28]). Another notable problem associated with using the LEIE dataset is that many of the providers listed in LEIE were prosecuted due to overt and deliberate fraudulent billing—their convictions were a diverse array of felonies such as billing under the National Provider Identifier (NPI) of another doctor, etc. The vast majority of healthcare abuse is attributed to overutilization. The subtle variations in billing patterns, which may show higher utilization of certain procedures and services than medically necessary is often undetectable by the non-physician. Using LEIE providers and their Medicare billing data as a training set may develop a model that can detect brazen, outlandish billing patterns, but would be unable to pinpoint instances of more subtle fraud from which a majority of the financial loss and waste occurs.

Another common issue observed in many Medicare fraud detection studies is the tendency to focus on aggregate information such as the total number of procedures performed, total Medicare reimbursement amount, etc. to detect fraudulent activity (Herland et al. [15]; Yao et al. [27]; Mayaki and Riveill [29]). While these aggregate features can provide a high-level overview, they may overlook crucial details about the specific procedures and services performed by the providers. By neglecting the granular information associated with each individual procedure, the model may miss out on identifying specific patterns or anomalies that could be indicative of fraudulent behavior. It is worth mentioning that one approach used to get around the data aggregation challenge is to assign overutilization labels to provider-procedure pairs instead of just the providers. This approach reflects the fact that fraudulent providers do not necessarily engage in fraudulent activities for every single procedure they perform. Bauder and Khoshgoftaar [12], Johnson and Khoshgoftaar [18], and Hancock and Khoshgoftaar [21] use the LEIE dataset to label the provider-procedure pairs. The issue with using the LEIE dataset is that it does not contain data on which procedures the provider submitted fraudulent claims for, and therefore all the provider-procedure pairs for the providers listed on the LEIE dataset should be labeled as fraud. This clearly could result in incorrect labels and introducing noise and bias into the training data, which in turn could significantly compromise the performance of the ML models. The substantial financial losses from fraud and abuse call for efficient analytical methods of fraud detection, involving both physicians and data scientists.

In this study, our primary goals are twofold. First, we aim to address the limitations of the LEIE dataset by incorporating the experience of seasoned physicians and medical billers to create a labeled dataset that overcomes the issues of class imbalance and the exclusive focus on overtly fraudulent providers. Unlike previous approaches our methodology involves capturing the nuances and intricacies of fraudulent behavior by incorporating the specific details of each procedure and service. By focusing our efforts on the field of ophthalmology, we leverage our access to domain knowledge and concentrate on the unique characteristics and specific Healthcare Common Procedure Coding System (HCPCS) codes relevant to this medical specialty. This targeted approach enables us to build a specialized dataset that reflects the intricacies and complexities of Medicare overutilization within ophthalmology.

Second, we conduct a comparative study of various machine learning models for the task of predicting Medicare overutilization within ophthalmology. To this end, we compare the performance of several supervised ML models including k-nearest neighbor, logistic regression, support vector machines, extreme gradient boosting, multilayer perceptron, as well as a stacking ensemble model. The comparative study not only enables us to identify the best-performing model but also aims to demonstrate the effectiveness of ensemble models in surpassing individual models in Medicare overutilization detection. Once the best-performing model is determined, we extend our analysis beyond model performance to estimate essential overutilization statistics and see which codes are most overutilized within ophthalmology. The estimated statistics include the overall overutilization rate within the specialty and the financial losses incurred as a result of these activities.

## 2 Methods

### 2.1 Data Source

The unprocessed data, without labels, is sourced from the CMS website [37], which currently encompasses data for 2013 to 2021 calendar years. Our analysis mainly relies on the Medicare Provider Utilization and Payment Data (MPUPD). [38] MPUPD comprises publicly available data files summarizing the information on services and procedures provided to Medicare beneficiaries by physicians and other healthcare professionals. Within MPUPD, we utilize two specific data files:

- Medicare Physician and Other Practitioners - by Provider and Service [40]: This data, referred to as MPOP PS, provides detailed information on use, payments, and submitted charges organized by NPI, HCPCS code, and place of service.
- Medicare Physician and Other Practitioners - by Provider [39]: This data, referred to as MPOP P, provides summary information on use, payments, submitted charges, and beneficiary demographic and health characteristics organized by NPI.

Note that both MPOP PS and MPOP P comprise individual datasets corresponding to each calendar year from 2013 to 2021. It is also worth mentioning that the data extraction process is specifically tailored to ophthalmologists, as our study focuses exclusively on this medical specialty. Table 1 summarizes the number of ophthalmology records found in MPOP PS and MPOP P in different calendar years. Appendix A provides further details on the data source.

**Table 1:** Number of ophthalmology records.

| Year | 2013 | 2014 | 2015 | 2016 | 2017 | 2018 | 2019 | 2020 | 2021 |
| --- | --- | --- | --- | --- | --- | --- | --- | --- | --- |
| MPOP_PS | 237,630 | 238,242 | 240,279 | 242,748 | 240,830 | 239,707 | 239,906 | 219,717 | 230,653 |
| MPOP_P | 17,550 | 17,664 | 17,698 | 17,788 | 17,817 | 17,804 | 17,856 | 17,631 | 17,489 |

### 2.2 Data Preprocessing

In our study, specific data preprocessing steps are taken to transform MPOP PS and MPOP P into a structured feature representation that captures the details of procedures and services for each provider in a given calendar year. The flowchart in Figure 1 illustrates a concise representation of the various stages involved in the data preprocessing step. In particular,

- Pivoting MPOP PS: We pivot the records based on the providers’ NPI to create a feature vector for each provider. This way, for each provider, we have a unique record detailing the procedures and services they have performed.
- Feature extraction from MPOP PS: In order to ensure a unified feature representation across all providers, we adopt a standardized set of 513 HCPCS codes pertaining to ophthalmology. For each HCPCS code, we extract two essential billing statistics from the pivoted MPOP PS—the number of services (NoS) and the number of Medicare beneficiaries (NoMB). This results in a feature set of size 1026 for each provider. More details on the HCPCS codes used can be found in Appendix B.
- Feature extraction from MPOP P: We extract only 3 features for each provider from the MPOP P data—total count of Medicare beneficiaries served by each provider (TotPatient), total payment made by Medicare to each provider (TotPayment), and the total payment made by Medicare to each provider for drug services (DrugPayment).
- Feature transformation: To enhance the informativeness of the features, we introduce two new features per HCPCS code—the ratio of NoS to NoMB and the ratio of NoMB to TotPatient. Additionaly, we introduce two supplementary features to further enrich the feature set—the ratio of TotPayment to TotPatient and the ratio of DrugPayment to TotPayment.

**Figure 1:**
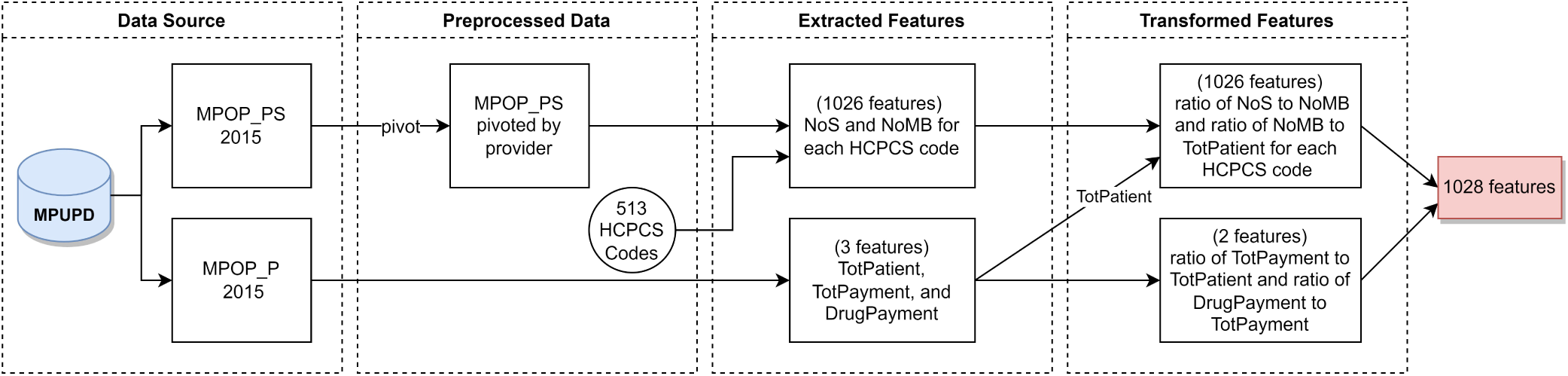
Data preprocessing flowchart. The presented flowchart outlines the sequential steps undertaken in the data preprocessing phase, specifically focusing on obtaining the feature set for a particular provider using their billing information from the year 2015.

By combining the transformed features, we obtain a feature set of size 1028 for each provider.

### 2.3 Dataset Labeling

Through collaboration with experienced physicians with a minimum of 10 years in practice and seasoned medical billers, we curated a labeled dataset consisting of 663 ophthalmologists based on their 2015 Medicare billing data.

The assigned labels are either “non-fraudulent” or “overutilizer”. The labeling process was conducted with the assumption that Medicare restrictions are consistent nationwide, ensuring equal opportunities for providers across the country to engage in overutilization activities. Each provider in the dataset was labeled by no less than 3 individuals to ensure consistency and accuracy. When there was any disagreement, an experienced fourth member of the team served as a tiebreaker to discuss and obtain consensus among the four. To ensure accuracy of labeling, further validation was made by a survey and test administered to practicing ophthalmologists. Baseline characteristics of the labeled providers can be found in Table 2. The labeling process is detailed in Appendix C.

**Table 2:** Characteristics of labeled providers.

| Characteristic | Overutilizer | Non-fraudulent |
| --- | --- | --- |
| Number | 200 | 463 |
| Location |  |  |
| California | 185 | 269 |
| New York | 38 | 171 |
| Average number of patients | 733.8 | 760.7 |
| Average number of services | 6904.3 | 4144.2 |
| Average Medicare payment per patient (\$) | 917.6 | 520.8 |

#### Preliminary statistical analysis

An initial examination of the generated features for the labeled providers highlights the significant sparsity (a large number of zero features) present in the feature set. Each provider is associated with 1028 features, out of which 1026 features are derived from 513 distinct HCPCS codes (2 features per code). However, not all the HCPCS codes are utilized by every provider, and on average, ophthalmologists bill for fewer than 50 HCPCS codes annually. For example, in 2015, ophthalmologists in California billed an average of just 33 HCPCS codes. Figure 2 illustrates the sparsity pattern observed in the features associated with the providers in the labeled dataset. The observed sparsity necessitates the use of special techniques during the training process. Appendix I further extends the analysis by comparing the generated features for the non-fraudulent providers vs that of the overutilizers aiming to identify any discernible difference between them.

**Figure 2:**
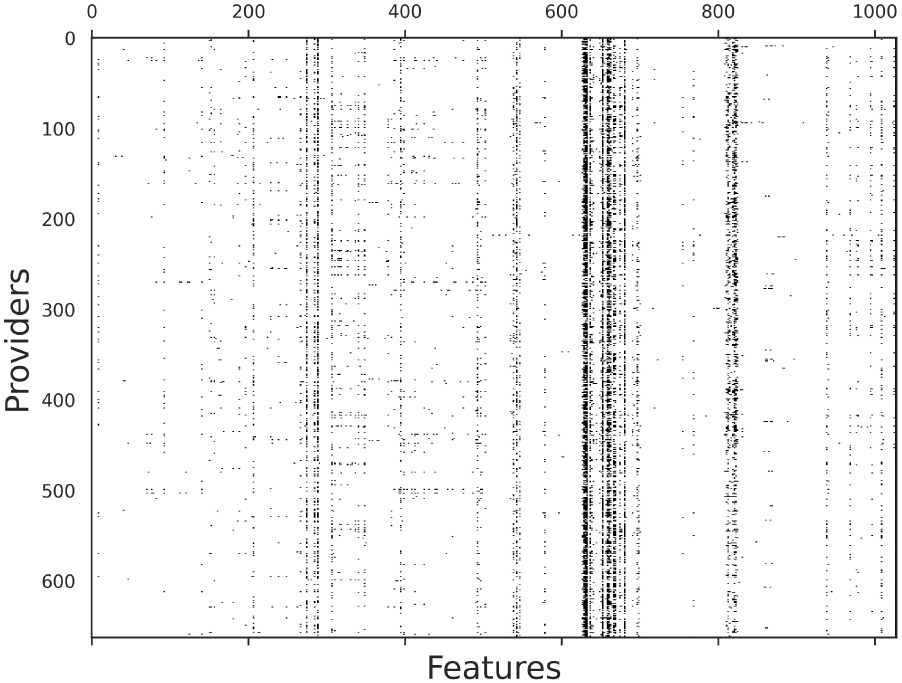
Sparsity map of the features associated with the providers in the labeled dataset. The plot shows the sparsity pattern of the feature space, indicating the utilization of HCPCS codes by labeled providers. Each cell represents the presence (non-zero value) or absence (zero value) of a specific HCPCS code for a provider.

## 3 Overutilization Analysis via Machine Learning Techniques

Providers’ billing behaviors, both the normal or the abnormal ones, can evolve over time due to various factors such as the introduction of new procedures, shifts in the medically necessary requirements of their patients, or efforts to avoid detection by auditors through normalized billing practices. To account for this dynamic nature, we employ models that aim to classify providers as either overutilizer (positive) or non-fraudulent (negative) based on their billing information within a specific calendar year. By focusing on yearly billing data, we can capture the evolving patterns and behaviors of providers, allowing for more accurate and timely detection of abnormal billing practice.

### ML models

In our analysis, we experiment with a diverse set of both linear and nonlinear predictive models including K-nearest neighbor (KNN), logistic regression (LR), support vector machines (SVM), extreme gradient boosting (XGB), and multilayer perceptron (MLP). To improve the model performance, we also explore an ensemble technique called stacked generalization (stacking) [1] to develop an ensemble model by combining the predictions of the five aforementioned ML models. The diverse capabilities of the individual models are shown to provide improved predictive accuracy. More details on the deployed ML models are provided in Appendix D.

### Performance evaluation

To ensure an unbiased evaluation of the models’ performance, a repeated nested cross-validation (nested CV) approach is employed. See Appendix E for detailed technical description. The models’ performance was evaluated using two metrics. The primary measure of performance is the Area Under the Receiver Operating Characteristic Curve (AUROC score), which quantifies the models’ ability to discriminate between positive and negative instances across different classification thresholds. Additionally, we report the Brier score assessing the models’ confidence by evaluating the accuracy of their probabilistic predictions.

We also plot Receiver Operating Characteristic (ROC) curves, which provide a graphical representation of the models’ discrimination ability across different classification thresholds. These curves allow us to compare the models’ performance in terms of true positive rate and false positive rate. Furthermore, we generate calibration plots to assess the calibration of the models. Calibration plots illustrate the agreement between the predicted probabilities of the models and the observed frequencies of the target variable. By visually examining the calibration plots, we can determine the models’ reliability and assess if they are underconfident or overconfident in their predictions.

### 3.1 Results

#### Model performance

The results of the model performance are presented in Table 3, which provides an overview of the mean performance scores along with their corresponding 95% confidence intervals (CIs) obtained through the repeated nested CV procedure. Additionally, Figure 3a displays the average ROC curves, showcasing the discriminative ability of each model.

**Figure 3:**
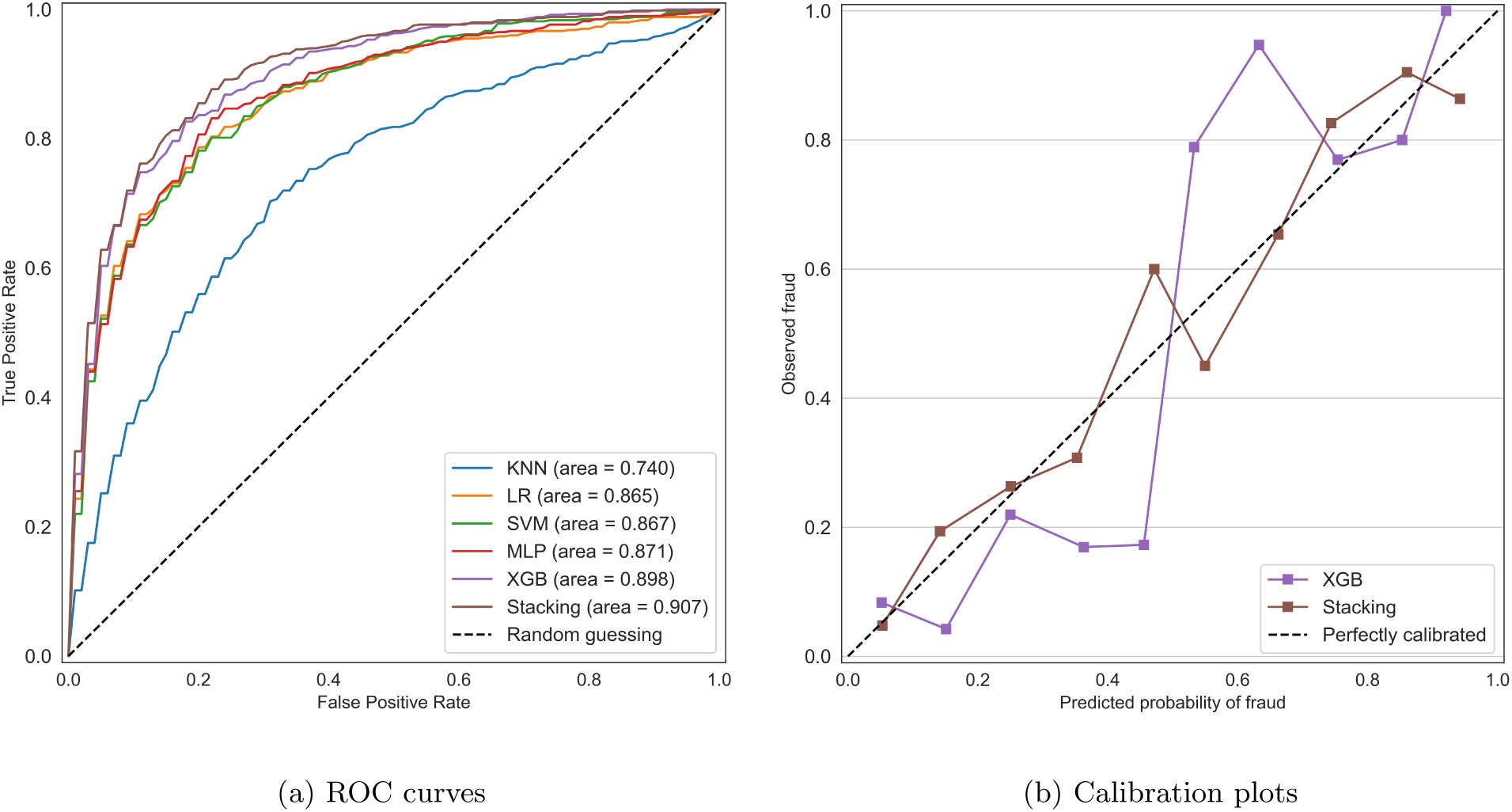
ROC curves and calibration plots. Corresponding values of the area under the curve (AUROC score) for each are presented in Table 3. KNN (blue line) indicates k-nearest neighbor; LR (orange line), logistic regression; SVM (greed line), support vector machines; MLP (red line), multilayer perceptron; XGB (purple line), extreme gradient boosting; and Stacking (brown line), stacking ensemble.

**Table 3:** Performance measures for various ML models.

| Model | AUROC score (95% CI) | Brier score (95% CI) |
| --- | --- | --- |
| KNN | 0.740 (0.716-0.765) | 0.182 (0.175-0.189) |
| LR | 0.865 (0.854-0.876) | 0.132 (0.127-0.138) |
| SVM | 0.867 (0.855-0.879) | 0.130 (0.123-0.137) |
| MLP | 0.871 (0.857-0.885) | 0.134 (0.125-0.143) |
| XGB | 0.898 (0.887-0.909) | 0.152 (0.136-0.168) |
| Stacking | 0.907 (0.896-0.918) | 0.109 (0.102-0.117) |

It is evident that all models exhibit a certain albeit varying level of discriminative power, as their mean AUROC scores surpassed that of a completely random classifier (AUROC score of 0.50). Among the models, the KNN model demonstrated the lowest discrimination with an AUROC score of 0.740 (95% CI: 0.716-0.765), indicating its limitations in accurately distinguishing between overutilizer and non-fraudulent instances. Meanwhile, the LR, SVM, and MLP models exhibited almost similar performance across all evaluation metrics, with strong discriminatory capabilities, achieving AUROC scores higher than 0.85. The XGB model displayed even better discriminative power, with an AUROC score of 0.898 (95% CI: 0.887-0.909). However, the higher AUROC score was accompanied by lower predictive accuracy, as indicated by the Brier score. In contrast, the stacking ensemble model demonstrated the strongest discriminative power, with an AUROC score of 0.907 (95% CI: 0.896-0.918), and the highest predictive accuracy and confidence.

The calibration plots in Figure 3b illustrate the consistency between the observed and predicted probabilities of overutilization for the XGB and stacking ensemble models. See Appendix F for the rest of the models’ calibration plots. Although the XGB model and the stacking ensemble model exhibit almost similar discriminative power (in terms of the AUROC score), a comparison of their calibration plots revealed a notable difference. The XGB model demonstrates lower consistency in its overutilization detection compared to the stacking ensemble model. Specifically, the XGB model is overconfident in its prediction, underestimating overutilization when the predicted probabilities are below 0.5 and overestimating overutilization when the predicted probabilities exceed 0.5.

### 3.2 Predicting Overutilization Statistics

Based on the model performance results, the stacking ensemble model is chosen and then trained on the entire labeled dataset to predict the labels for all ophthalmologists across the United States based on their Medicare billing data from 2021. Consequently, various overutilization statistics, including the Medicare overutilization rate and the monetary losses attributed to Medicare overutilization within the field of ophthalmology are obtained. We first conduct these calculations across the entire nation, and then within individual Medicare jurisdictions. Furthermore, we present a heatmap depicting the predicted overutilization rates across different states in the United States.

#### Nationwide overutilization rate

Among the 17,013 ophthalmologists enrolled in Medicare in 2021, our analysis predicts that 1,457 of them are likely to engage in overutilization activities, resulting in a predicted overutilization rate of approximately 8.6%.

#### Nationwide monetary loss

We compute the average Medicare payment per patient (MPPP) for both the predicted overutilizer and non-fraudulent providers. The 1,457 overutilizer ophthalmologists (mean=$908.0, std=$1021.2) compared to 15,556 non-fraudulent ophthalmologists (mean=$542.0, std=$841.3) had significantly higher MPPP (t(17011)=13.3, p *<<* 0.01). The t-test is conducted using Scipy v1.8.1 [24]. This suggests that the overutilizer ophthalmologists tend to receive higher Medicare payments per patient compared to their non-fraudulent counterparts. Additionally, these overutilizer providers had a combined patient count of 932,520 and the total Medicare payment to them amounted to $942.5 million. To put these numbers into perspective, if all the overutilizer physicians had similar MPPP to the average non-fraudulent physician ($542.0), their total Medicare payment would have been $505.4 million (932, 520 *×* 542.0). This indicates that there is a potential loss of $437.1 million due to overutilization activities for just one year and just the field of ophthalmology.

We further provide a breakdown of the calculated statistics across different Medicare jurisdictions and states. Table 4 provides an overview of the 12 Medicare jurisdictions, including the states and territories they cover. The calculated statistics within each Medicare jurisdiction are shown in Figures 4 and 5. Figure 6 depicts the heat map of overutilization rates across different states in the US. More details on the Medicare jurisdictions’ overutilization statistics are provided in Appendix G.

**Figure 4:**
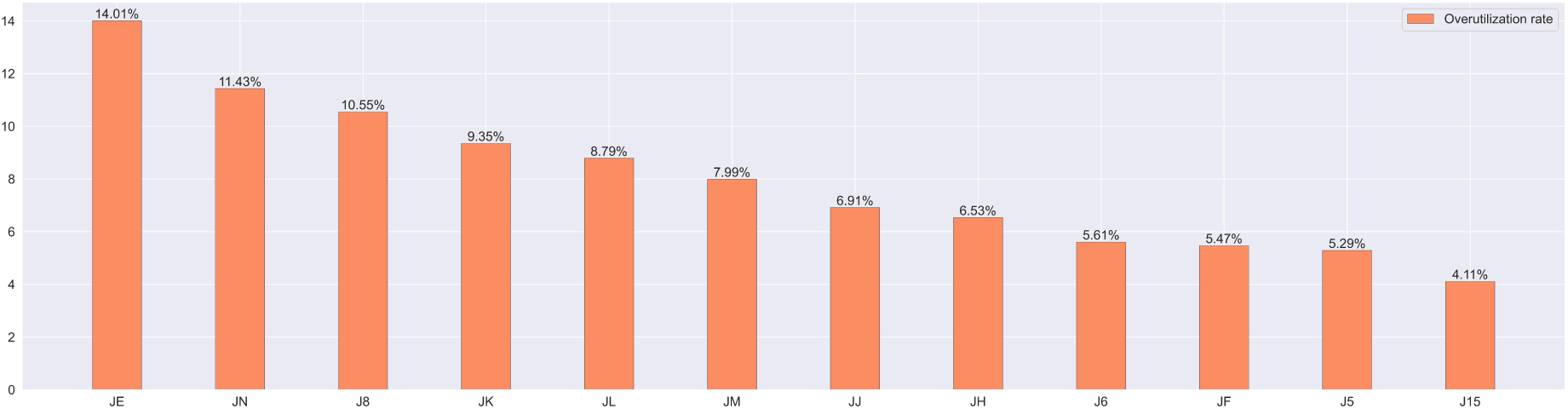
Predicted Overutilization Rates within Medicare Jurisdictions in 2021. The figure displays the overutilization rates, sorted in descending order, for each Medicare jurisdiction. The leftmost jurisdiction represents the highest overutilization rate, while the rightmost jurisdiction indicates the lowest fraud rate. The data provides insights into the distribution of predicted overutilization across different jurisdictions.

**Figure 5:**
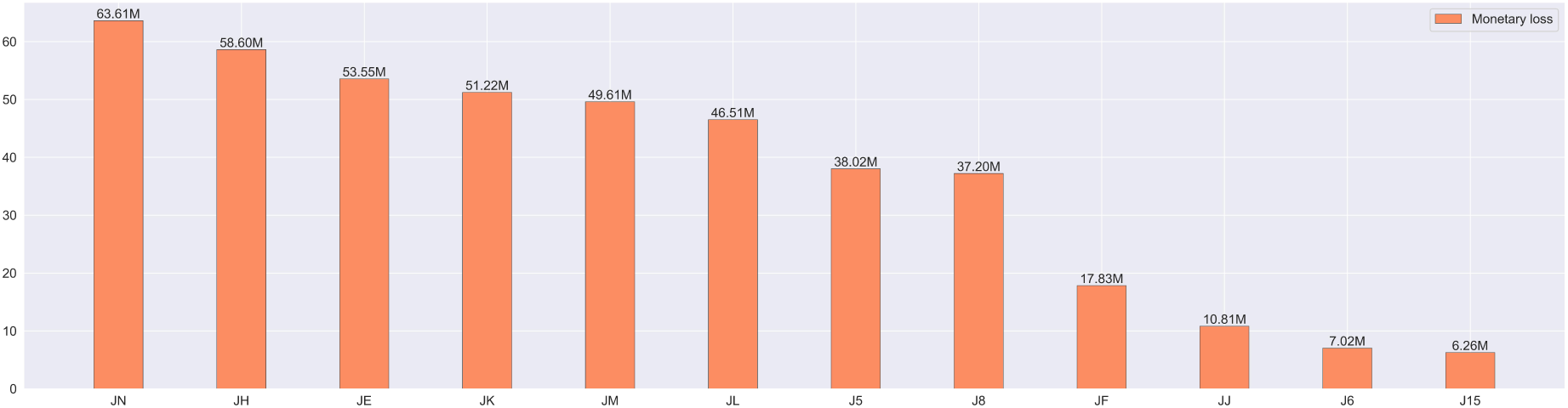
Monetary Losses on Medicare overutilization within Medicare Jurisdictions in 2021. The figure illustrates the amount of money lost due to overutilization, sorted in descending order, for each Medicare jurisdiction. The leftmost jurisdiction represents the highest monetary losses, while the rightmost jurisdiction indicates the lowest losses. The losses are calculated in the same way nationwide loss is calculated.

**Figure 6:**
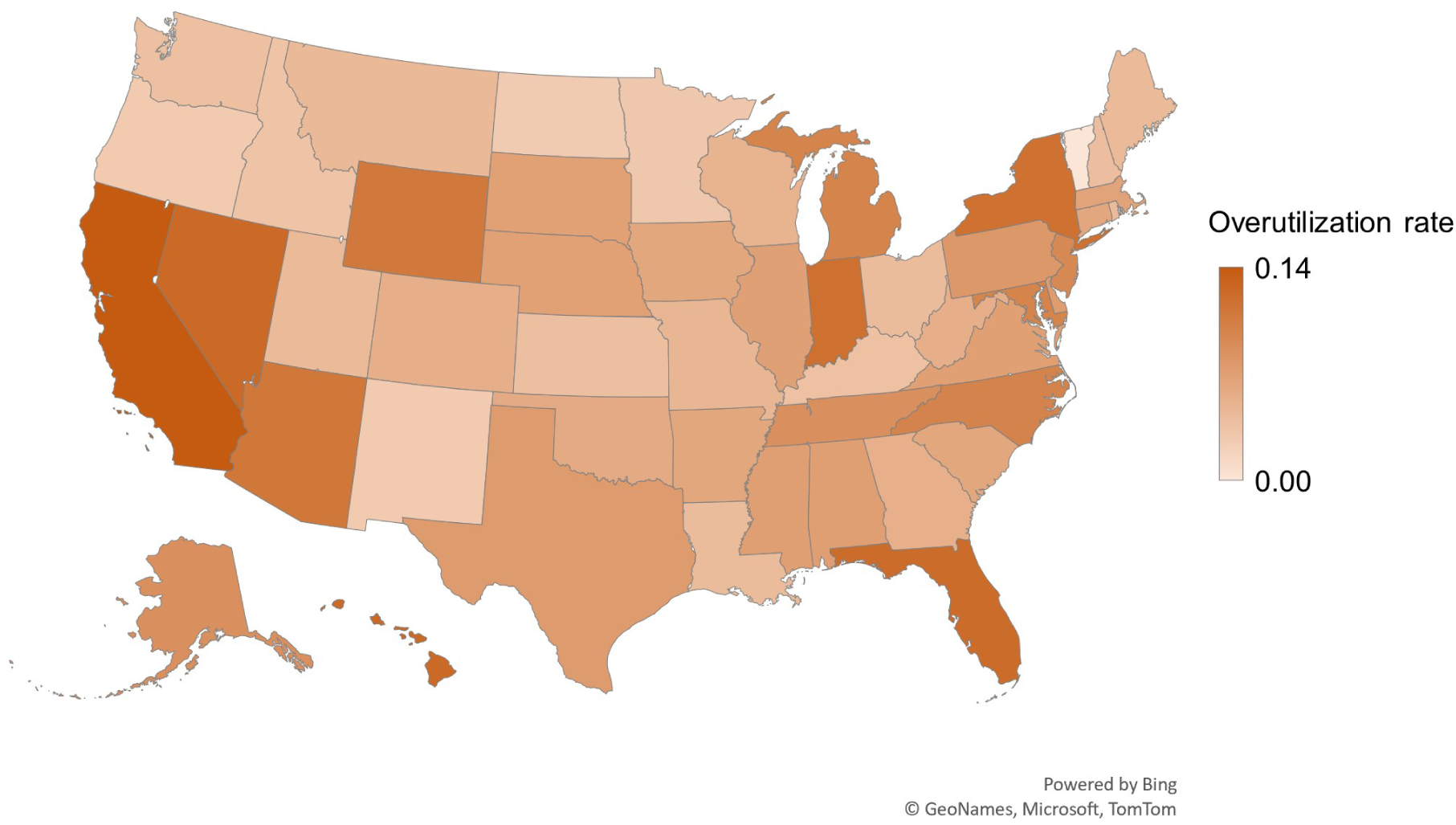
The heat map showcases the predicted overutilization rates across states in the United States, providing a visual representation of the variations in overutilization patterns across different regions. The darker colors represent higher rates of overutilization.

**Table 4:** Medicare jurisdictions and coverage areas.

| Jurisdiction | States and territories | Number of ophthalmologists in 2021 |
| --- | --- | --- |
| JE | CA, NV, HI, AS, GU, MP | 2149 |
| JF | WA, OR, ID, MT, WY, ND, SD, UT, AZ, AK | 1243 |
| JH | CO, NM, TX, OK, AR, MS, LA | 2143 |
| JL | PA, MD, DE, NJ, DC | 1855 |
| J5 | NE, KS, IA, MO | 662 |
| J8 | MI, IN | 825 |
| JJ | TN, AL, GA | 926 |
| JM | WV, VA, NC, SC | 1264 |
| JK | NY, VT, MA, CT, RI, NH, ME | 2502 |
| J6 | MN, WI, IL | 1248 |
| J15 | OH, KY | 779 |
| JN | FL, PR, VI | 1417 |

We remark that the significant variation in MPPP, as indicated by the high standard deviation calculated earlier, is primarily attributed to the inclusion of drug payments. If we exclude drug payments from the total payments, we anticipate that the average payment per patient for non-drug services among overutilizer providers will still be considerably higher than that of non-fraudulent providers. See Appendix G for a comparison.

It is also worth mentioning that in order to obtain precise labels for overutilization detection, we employed a specific probability threshold to convert the predicted probabilities into suspected overutilizer versus non-fraudulent labels. In order to strike a balance between precision and sensitivity (recall), we select a probability threshold of 0.353. With this threshold, the stacking ensemble model achieves an accuracy of 0.850 (95% CI: 0.836-0.862), a specificity of 0.886 (95% CI: 0.867-0.903), a precision of 0.750 (95% CI: 0.720-0.779), and a sensitivity of 0.763 (95% CI: 0.733-0.793). More information on selecting the probability threshold is provided in Appendix H.

### 3.3 Feature Importance Analysis

Feature importance analysis is a valuable technique in ML that enables understanding the relative importance of different features in making predictions. By assessing the impact of features on the stacking ensemble model’s output, we can gain insights into which factors play a significant role in influencing the model’s decisions in the detection of overutilization. One popular approach to feature importance analysis is using SHAP (SHapley Additive exPlanations).[11] SHAP quantifies the contribution of each feature in the prediction process. We utilize the Python package SHAP v0.41.0 [11] in our study.

Figure 7 shows the SHAP summary plot to visualize the feature importance values obtained from the stacking ensemble model analysis. In this plot, the features are listed along the y-axis of the plot, with the most important features at the top. Secondly, each feature is represented by a horizontal bar in the plot depicting the corresponding SHAP values. A positive SHAP value means positive impact on prediction (in our case, overutilization/fraud prediction), whereas a negative SHAP value corresponds to a negative impact. A higher positive value indicates a higher positive impact, and a lower negative value indicates a higher negative impact. Lastly, the color gradient within each bar represents the value of the corresponding feature.

**Figure 7:**
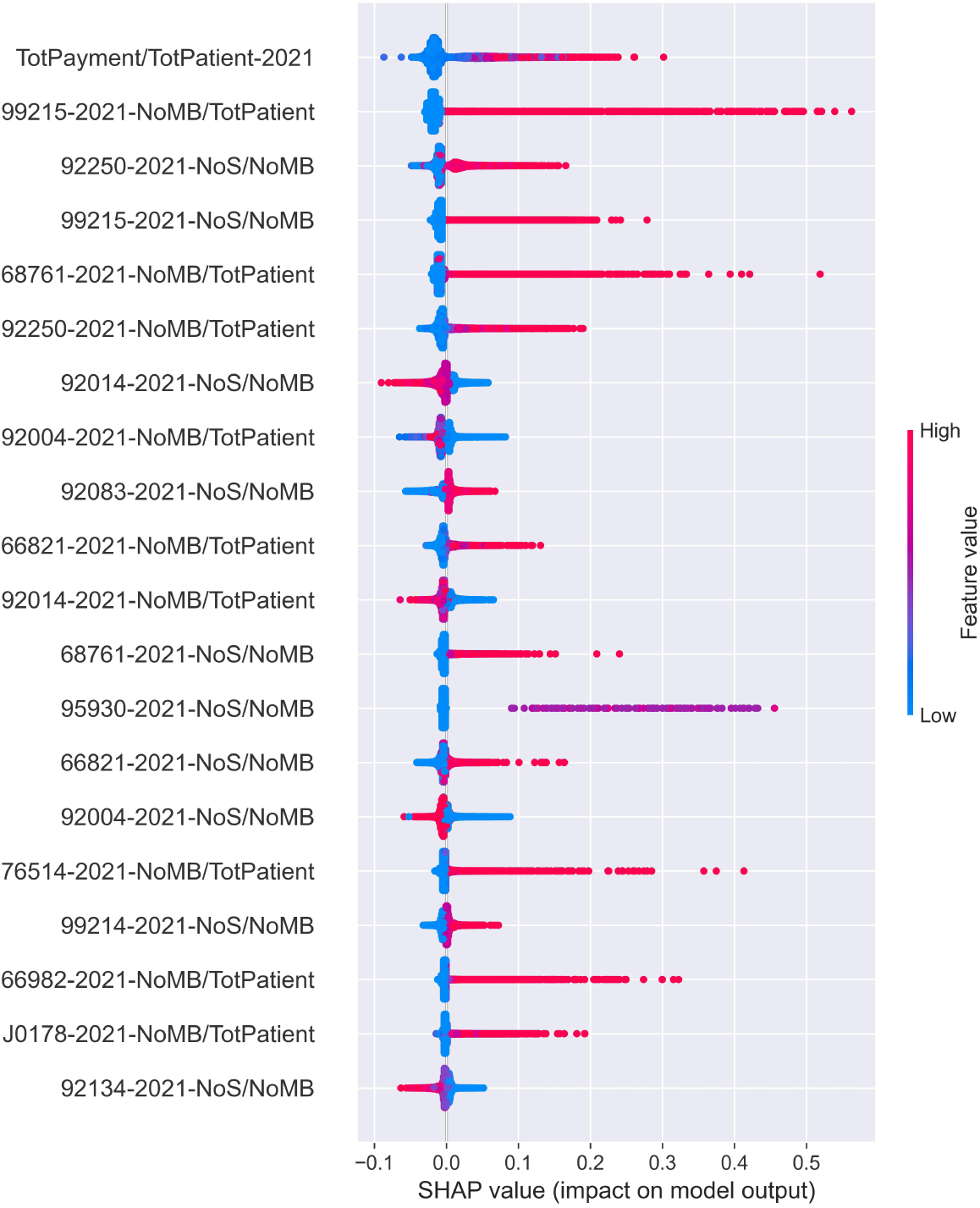
Shap Summary Plot.

The first feature displayed at the top is the ratio of total amounts of Medicare payments to the total number of patients. The corresponding horizontal bar suggests that higher values of this ratio consistently have a positive impact on the model’s predictions of overutilization. Other features listed in the plot present the HCPCS codes that could be more influential in determining overutilization.

## 4 Discussion

The goal of our study was to test various ML models to identify the most effective screening approach for Medicare overutilization detection within the field of ophthalmology. This was uniquely accomplished by curating a comprehensive labeled dataset of ophthalmologists. Our medical team ensured the labeled dataset addressed the limitations of the LEIE dataset by tackling the issue of class imbalance and capturing nuanced fraudulent patterns and overutilization. Using our created dataset, we found that the stacking ensemble model enhanced overutilization detection by harnessing the strengths and diverse perspectives of individual ML models. In what follows, we further discuss the different components of our analysis and the key findings.

The engineered features are designed in very labor-intensive collaboration with physicians and medical experts, ensuring that they align with their domain expertise and thought process during the labeling of the data. First, the ratio of NoS to NoMB provides insights into the frequency of services rendered per beneficiary, allowing for a more nuanced understanding of utilization patterns. Second, the ratio of NoMB to TotPatient provides valuable information about the proportion of Medicare beneficiaries among all patients served, which can shed light on the demographics and patient population of the provider. Additionally, the ratio of TotPayment to TotPatient reflects the average Medicare payment per patient, offering insights into the financial aspects of the services provided. Lastly, the ratio of DrugPayment to TotPayment captures the proportion of drug-related payments within the total Medicare payment amount.

To evaluate the effectiveness of these features in overutilization detection, consider the following scenarios. Take the HCPCS code 66984, which represents extracapsular cataract removal. If the ratio of the NoS to NoMB ratio exceeds two in a given year, it would be highly suspicious. This is because, on average, each patient has only two eyes, and the average number of cataract surgeries per patient should not exceed two. (Note that the revision surgery has a separate code and can be distinguished from initial surgery.) Similarly, for the HCPCS code 95004, which pertains to allergy testing of the skin, if the NoMB to TotPatient ratio is 1 or higher, it indicates that allergy testing has been performed on every patient encounter. In the case of ophthalmologists, this suggests excessive and unwarranted billing since not every patient requires skin allergy testing every encounter. Additionally, the ratio of DrugPayment to TotPatient is particularly useful in distinguishing retina specialists from other subspecialties. The aforementioned ratio is often high among retina specialists due to the expensive nature of the anti-VEGF drugs they utilize.

Our labeled dataset provides a more balanced distribution between overutilizer and non-fraudulent providers compared to the LEIE dataset, with the minority class representing 30.2% of the dataset. In the specific context of ophthalmology, the LEIE dataset includes only 63 fraudulent ophthalmologists, while in 2021, there were over 17,000 ophthalmologists participating in Medicare. It is important to note that the LEIE dataset primarily focuses on extreme cases of fraud and may not capture the full range of overutilizing activities in the field. In contrast, our collaborative approach with physicians and medical billers allowed us to thoroughly examine the billing information of selected ophthalmologists, ensuring that our dataset represents a wider range of overutilizer and non-fraudulent providers. The detailed examination of billing patterns can only be accomplished when physicians or medical billers with years of experience are analyzing the data. Unfortunately this has never been done before because of the labor intensive nature of creating the dataset as well as industry wide lack of domain expertise, i.e., claim analysis is rarely done by physicians and those who understand the patterns of clinical care.

The comparative evaluation of different ML models revealed varying levels of discrimination performance in detecting Medicare overutilization within the field of ophthalmology. The KNN model exhibited the lowest discrimination ability, while the LR, SVM, and MLP models demonstrated similar strong performance. The XGB model displayed even better discrimination power, although with lower predictive accuracy. However, it was the stacking ensemble model that consistently showcased the highest performance. The superior discrimination performance as well as the high predictive confidence of the stacking ensemble model indicates that combining the predictions of multiple models using the stacked generalization technique can lead to improved overutilization detection outcomes. Therefore, more attention should be directed towards ensemble approaches, as they have the capability to leverage the strengths of individual models and produce improved overutilization detection outcomes.

Using our stacking ensemble model trained on the entire labeled dataset, we were able to estimate the nationwide overutilization rate as 8.6%, which falls within the range of existing fraud estimates of 3-10%.[5] Additionally, we estimated the annual monetary loss on overutilization and fraud in Medicare for the field of ophthalmology to be approximately $437 million. By extrapolating this estimate to the entire Medicare system, considering ophthalmology’s share of 0.89% of the Medicare budget, we estimate the total money lost on overutilization and fraud in Medicare to be around $49.1 billion per year. Considering the $900.8 billion total cost of Medicare program in 2021, this implies that approximately 5.5% of the Medicare budget is lost due to overutilization and possible fraud. These findings highlight the dire need for effective prevention strategies.

Current prevention and detection strategies prove to be very insufficient. CMS often outsources and relies on overly simplistic ratios to identify potentially irregular behavior. For example, one analysis is the ratio of complex cataract surgery to total surgeries done by a physician. This would not necessarily account for those physicians expert in complex cataract surgery who are referred these particularly difficult cases. Additionally, these analyses lack the consideration of subspecialties and fail to capture the intricacies and complexities of physician billing practices. Incorporating real-time machine learning for claims analysis would be a huge step forward in both the deterrence and post claim payment recoupment of overutilization. This way, flagged claims could undergo further scrutiny without the need to manually review thousands of appropriately billed and paid claims

We additionally conducted estimations of overutilization rates and associated monetary losses within each Medicare jurisdiction. The predicted rates exhibited variations across different regions of the United States. Notably, Jurisdiction JE, which includes California, displayed the highest predicted overutilization rate at 14.01%, followed by JN with 11.43%. Conversely, the jurisdiction J15 encompassing Ohio and Kentucky had the lowest overutilization rate at 4.11%. Surprisingly, although jurisdiction JE had the highest overutilization rate, it did not have the highest monetary loss. Instead, JN incurred the highest monetary loss at $63.6 million, followed by JH with $58.6 million. Several factors could contribute to this distribution, including demographic disparities, variations in healthcare provider density, regional differences in fraud awareness and enforcement, cultural and behavioral norms, and variances in healthcare practices. These findings underscore the need for targeted measures for detecting and preventing overutilization that are customized to specific regions.

The feature importance analysis revealed that the payment per patient ratio was a strong indicator of overutilization, with higher values indicating a higher likelihood of overutilization activity. This association can be attributed to the potential incentive of using certain highly reimbursed HCPCS codes frequently, even when not medically necessary. Moreover, the excessive utilization of specific HCPCS codes has been identified as a significant indicator of overutilization. This heightened utilization is observed through either a high ratio of beneficiaries to total patients or a high ratio of services to beneficiaries. The HCPCS code 95930 (visual evoked potential) is an example to demonstrate how excessive use serves as a red flag for fraudulent behavior. In ophthalmology, this code is narrowly applicable, primarily employed by neuroophthalmologists to diagnose optic neuropathies or malingering through measurements of the visual cortex’s response to visual stimuli. However, when a general ophthalmologist utilizes this code for nearly every patient and every visit, it deviates from the standard of care and signifies excessive utilization. It is apparent that there may be a financial motivation behind this utilization. It is noteworthy that the Medicare fee schedule allowable for HCPCS code 95930 is $134.35 in certain geographic locations, and when multiplied by all the patients in a given practice can be financially very lucrative.

It is crucial to mention that the high utilization of an HCPCS codes is not necessarily indicative of fraud. For instance, an ophthalmologist may have a very high usage of codes such as 99215 (highest reimbursing patient examination code) as well as many esoteric codes such as the aforementioned 95930 or 92275 (retinal electrography). Individual code analysis may pinpoint this physician with overutilization, but an analysis of all claims of their practice may reveal that he or she has a neuro-ophthalmology subspecialty, with very complex patients and few total encounters over the course of a year compared to a high volume general ophthalmologist. This suggests that overutilization detection cannot solely rely on individual code analysis but should consider patterns and relationships among multiple codes to gain a comprehensive understanding of potential fraudulent behavior.

Never before has domain expertise of ophthalmologists and medical billers been combined with machine learning to analyze the medicare claims database. The implications of this study are that medicare could save $50 billlion dollars per year by incorporating a properly trained machine learning model under each specialty in its claims analysis. We focused on the most egregious and blatant forms of overutilization. If applied to more subtle forms of overutilization, then the cost savings would be even greater. The additional funds generated could be applied to lowering the eligibility age of medicare recipients, expanding clinical and drug coverage, or raising reimbursement rates for the vast majority of honest physicians providing clinically and epidemiologically appropriate care. In this modern era where machine learning has the power to analyze individual provider behavior, and to compare it with the totality of the medicare data set, it is long overdue that we leverage the computational power of big data, to use medicare dollars efficiently and appropriately.

## 5 Limitations

Despite the aforementioned strengths and critical findings, our study has a number of limitations. The first limitation of our work is the limited number of data points in our created dataset. The meticulous and labor intensive process of analyzing and evaluating billing information for each provider resulted in a relatively small dataset. This limited sample size may not capture the full variations and distributions of overutilizer and non-fraudulent data, particularly given its high-dimensional nature. To enhance the accuracy and generalizability of the models, further data acquisition efforts are necessary to expand the dataset.

Secondly, it is important to acknowledge that the reported overutilization statistics are based on model predictions, which are not infallible. As such, it is crucial to interpret the model’s predictions as estimations rather than absolute truths. Validation and verification through additional means, such as manual audits or investigations, are necessary to ascertain the true extent of overutilization and possibly fraudulent activities, and the associated financial losses. While the findings can guide decision-making and resource allocation, ongoing refinement and improvement of the models and methodologies are essential to ensure their accuracy and reliability in detecting aberrant behavior.

Further, it is often difficult to distinguish high or overutilization from blatant fraud, which has intent behind it. Utilization ultimately is a broad continuum and often the medical record is needed to justify clinical behavior. This paper addresses clinician behavior by yearly patterns, but Medicare ascertains fraud by individual patient records to determine if a procedure was done and if clinically appropriate. Often on an individual basis, actions can be defensible, even if occurring on a repeated basis.

Lastly, it is worth noting that although some components of our study, such as data preprocessing and feature engineering, can be applied to detect Medicare overutilization in other subspecialties, the availability of a labeled dataset specific to each subspecialty remains a requirement. This underscores the need for domain expertise and collaboration with healthcare professionals in that subspecialty to create labeled datasets that accurately capture overutilization patterns within specific subspecialties. This can be very labor intensive and costly and represents the largest barrier to widespread incorporation of this methodology.

## Data Availability

All data produced in the present study are available upon reasonable request to the authors

## Appendix A

### MPOP_PS data

This data is structured in comma-separated values (CSV) format and comprises individual files for each calendar year. Every record in the data identifies a provider, primarily by the provider’s National Provider Identifier (NPI), and a procedure that the provider has submitted a claim to Medicare for, which is represented with a Healthcare Common Procedure Coding System (HCPCS) code. In addition to the NPI, each record contains several other elements that offer detailed information about the provider including their name, demographics, and the provider’s type (e.g. ophthalmology). Furthermore, each record includes essential aggregate statistics pertinent to the listed procedure. These statistics encompass the number of times the provider performed the procedure within the given year, the average billing amount for that procedure, and other relevant information. Table 5 lists the subset of features from the MPOP PS data used in our analysis.

**Table 5:** Features used from the MPOP_PS data.

| Name | Description | Type |
| --- | --- | --- |
| Rndrng_NPI | National Provider Identifier (NPI) for the rendering provider on the claim | Categorical |
| HCPCS_Cd | HCPCS code used to identify the specific medical service furnished by the provider | Categorical |
| Tot_Srvcs | Number of services provided | Numerical |
| Tot_Benes | Number of distinct Medicare beneficiaries receiving the service | Numerical |

### MPOP_P data

This data is structured in comma-separated values (CSV) format and comprises individual files for each calendar year. Each record in the data represents a unique provider, primarily identified by their National Provider Identifier (NPI). Additional information provided by each record includes demographic details, provider’s type, and various aggregate statistics related to their services and procedures such as total number of unique HCPCS codes, total number of Medicare beneficiaries receiving services from the provider, etc. Table 6 lists the subset of features from the MPOP P data used in our analysis.

**Table 6:**
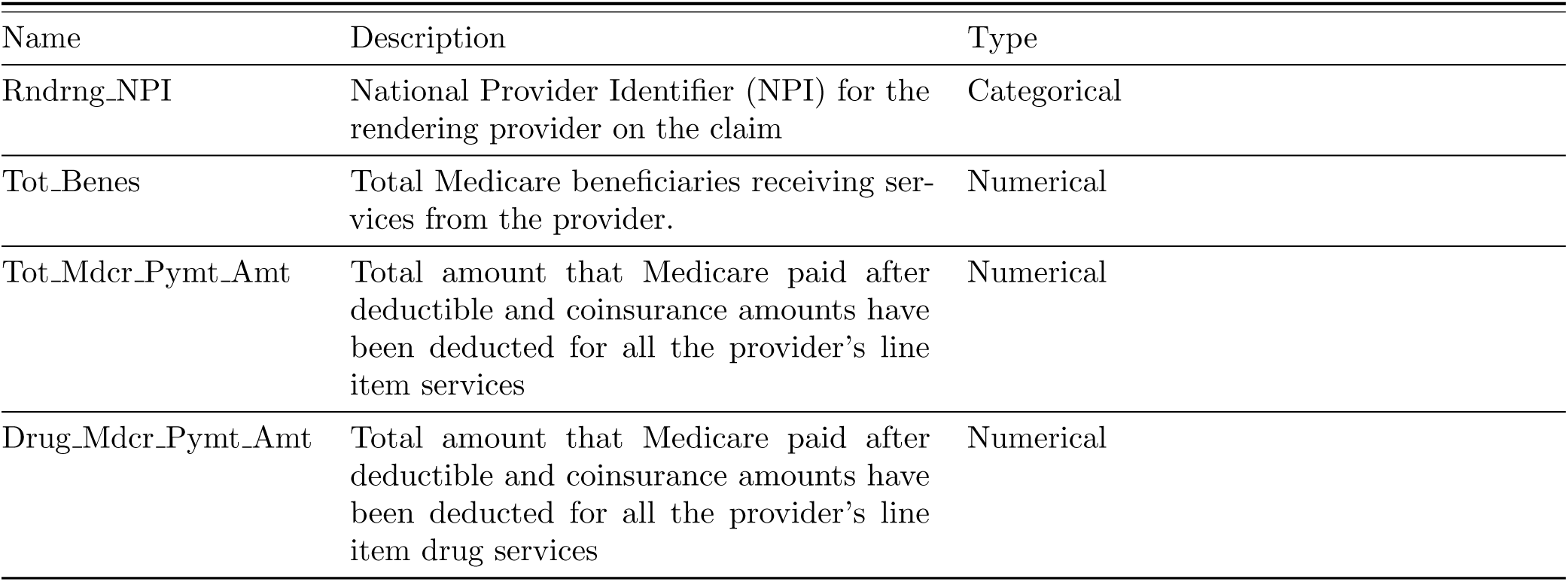
Features used from the MPOP_P data.

| Name | Description | Type |
| --- | --- | --- |
| Rndrng_NPI | National Provider Identifier (NPI) for the rendering provider on the claim | Categorical |
| Tot_Benes | Total Medicare beneficiaries receiving services from the provider. | Numerical |
| Tot_Mdcr_Pymt_Amt | Total amount that Medicare paid after deductible and coinsurance amounts have been deducted for all the provider’s line item services | Numerical |
| Drug_Mdcr_Pymt_Amt | Total amount that Medicare paid after deductible and coinsurance amounts have been deducted for all the provider’s line item drug services | Numerical |

## Appendix B

The set of 513 HCPCS codes used to construct the feature vectors is derived from aggregating the services and procedures performed by ophthalmologists located in California over the period from 2013 to 2019. This timeframe allows us to capture a substantial amount of data and encompass the diversity of services rendered by ophthalmologists during those years. “hcpcs codes.xltx” lists all the 513 HCPCS codes used.

California, being home to the highest number of ophthalmologists compared to other states (in 2015, 1,975 out of 17,698 ophthalmologists in the United States were located in California), provides a rich source of information for constructing a representative set of HCPCS codes. By leveraging the data from California, we can ensure that the selected 513 HCPCS codes encompass a wide range of procedures and services commonly performed within the field of ophthalmology.

## Appendix C

The initial set of labels was generated manually by utilizing the Wall Street Journal’s Medicare Unmasked graphic [41], which provided visual representations of Medicare reimbursements received by providers in a given year. This program offered insights into various aspects, including total payments, patient counts, payments per patient, and a breakdown of reimbursements by service category. It also provided information on HCPCS codes used, the number of unique patients receiving each procedure, and the total Medicare reimbursement for specific HCPCS codes in the year under consideration (2015 in our case). The labeling team thoroughly examined this information to determine the epidemiological accuracy of the billed HCPCS codes. This involved considering factors such as the ratio of HCPCS codes to the number of Medicare beneficiaries, the ratio of higher-reimbursing procedures to lower-reimbursing procedures, and the provider’s reimbursement percentile at the state and national levels.

The labeling process involved multiple iterations of training the SVM model on the already labeled dataset, using the trained model to predict the likelihood of unlabeled providers being fraudulent, and selecting a random sample of 50 providers who were on the borderline (with a probability close to 0.5) for further labeling by the team. Sampling providers who were on the borderline helped improve the model’s ability to distinguish between fraudulent and non-fraudulent cases. While it was relatively easier to identify blatantly fraudulent or non-fraudulent doctors, borderline cases posed greater challenges. Obtaining labels for this borderline sample contributed to enhancing the model’s accuracy in detecting fraud.

However, due to the difficulty in definitively determining the fraudulent or non-fraudulent nature of borderline doctors, the process of generating these labels was time-consuming. Manual analysis encompassing various dimensions, such as the number of services performed per code, the total number of patients for each physician, and the physician’s subspecialty, was conducted to create a label for each billing pattern. Given the meticulous nature of this analysis, generating an adequate number of labels to train the model required a significant amount of time

## Appendix D

### Models’ Configuration and Hyperparameters

In our analysis, we develop a diverse set of both linear and nonlinear predictive models including k-nearest neighbor (KNN), logistic regression (LR), support vector machines (SVM), extreme gradient boosting (XGB), and multilayer perceptron (MLP). Each model is embodied in a specific pipeline that incorporates various preprocessing techniques, such as feature scaling (e.g., min-max normalization, standardization) and dimensionality reduction methods (e.g., principal component analysis, recursive feature elimination, feature subsampling) to enhance their performance. It is important to note that when we refer to the models, we are specifically referring to the complete pipelines they are part of. Figure 8 shows the exact configuration and hyperparameters associated with the deployed pipelines.

**Figure 8:**
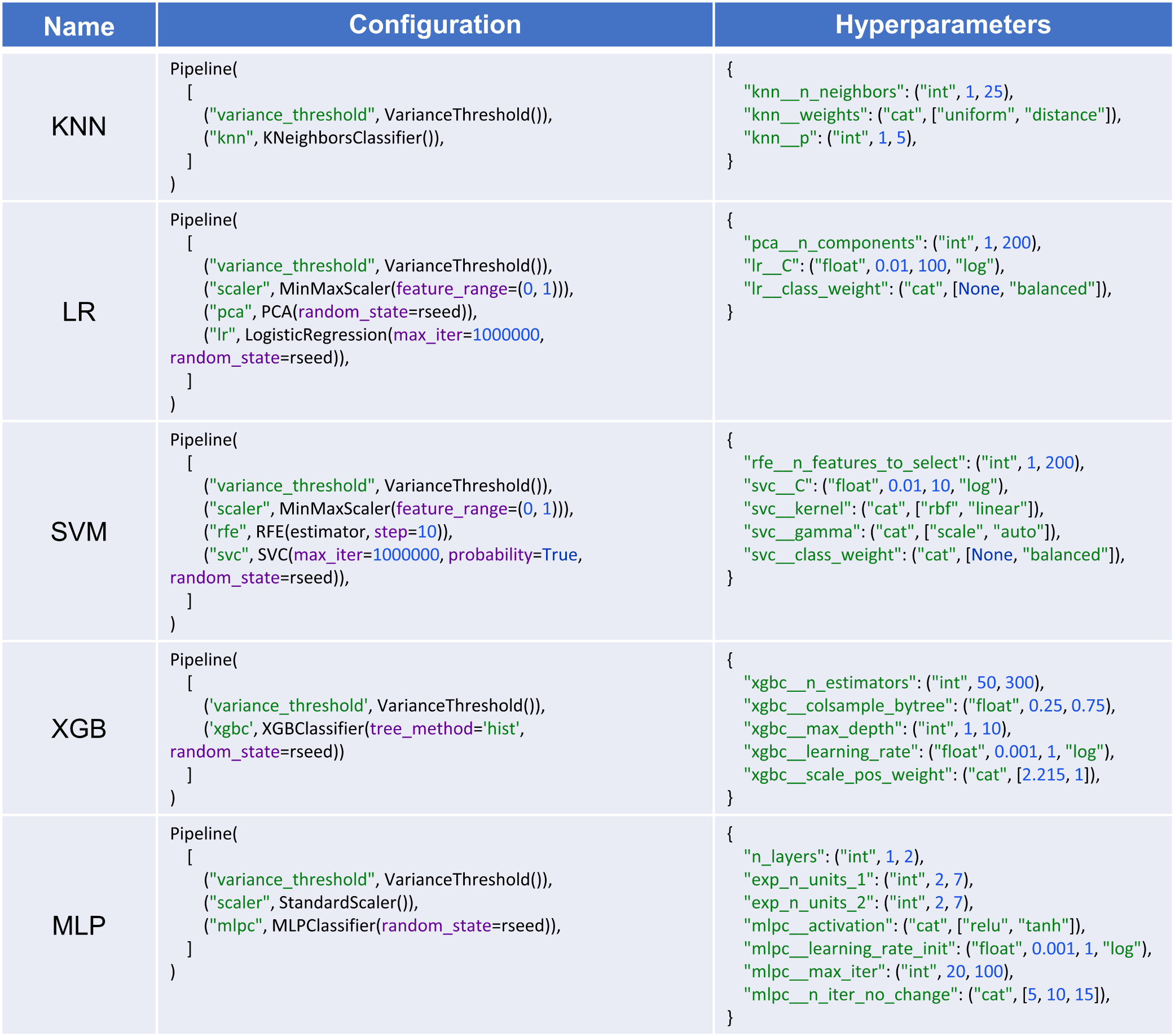
Configuration and hyperparameters of deployed pipelines. The figure illustrates the detailed configuration and hyperparameter settings for the pipelines utilized in the training process.

The hyperparameter settings are used during the nested CV procedure to fine-tune the models. For each model, the hyperparameter settings are defined as a Python dictionary in which each key defines a hyperparameter and its corresponding value determines the type and search domain for that hyperparameter. The type and range of a hyperparameter is formatted as a tuple in which the first element defines its type and could be either `“int”` (integer), `“float”`, or `“cat”` (categorical). The rest of elements determine the search domain. For instance, (`“int”, 1, 25`) defines an integer between 1-25, or (`“float”, 0.01, 10, “log”`) defines a float in the range 0.01-10 sampled in the logarithmic domain. Additionally, `(”cat“, [None, “balanced”])` determines a categorical variable which could be either `None or “balanced”`.

Further information regarding the pipelines’ configurations and hyperparameters are listed below:

- All analyses are done using Python v3.8. The KNN, LR, SVM,and MLP models were implemented using scikit-learn v1.1.2 [4]. The XGB model is implemented using XGBoost v1.6.2 [8].
- To expedite the training process, preprocessing methods like feature scaling are employed. The primary technique utilized for feature scaling is min-max normalization, which preserves the non-negativity and sparsity of the features. Standardization, on the other hand, is exclusively applied to MLP due to improved performance compared to min-max normalization, as observed in our experiments.
- In our analysis, we employ multiple techniques for dimensionality reduction. The Sklearn library’s VarianceThreshold module is utilized to eliminate constant features, which are essentially features that consist entirely of zeros. Additionally, PCA (Principal Component Analysis) and RFE (Recursive Feature Elimination) are specifically employed in the LR and SVM pipelines, respectively. Furthermore, in the XGB pipeline, we restrict the model from utilizing the entire feature set by setting the colsample_bytree argument to a value less than one.
- Certain models including LR, SVM, and XGB offer the ability to address class imbalance by resampling the training data. For instance, by setting `“lr class_weight” to “balanced”`, the LR model oversamples the minory (fraudulent) class. This helps mitigate the impact of class imbalance.

### Stacking

In addition to the individual models, we also explore the benefits of ensemble techniques in improving overall performance. We leverage stacked generalization (stacking) [1] to develop an ensemble model, which combines the predictions of the five aforementioned ML models. Stacking involves the use of an additional model called the “meta-learner” to learn the optimal way of combining the predictions from the base models. In our ensemble model, the base models, KNN, LR, SVM, XGB, and MLP, generate predictions that are then fused together using an LR-based meta-learner. This ensemble approach allows for leveraging the diverse capabilities of the individual models and can lead to improved predictive accuracy. The exact meta-learner used in our analysis is `LogisticRegression(penalty=“none”, max_iter=1000, random_state=0)`.

In the following explanation, we outline the training and testing process for the stacking ensemble model using a given training set. Initially, the 5 base models undergo fine-tuning through k-fold cross-validation on the training set. During this procedure, the predictions made by each tuned base model are stacked together. By merging the stacked predictions from all the tuned base models, we construct the training set for the meta learner. Subsequently, all the tuned models are trained using the entire training set, while the meta learner is trained using its corresponding training set. For testing and making predictions on unseen data, we first utilize the trained base models to generate predictions, and then these predictions are combined using the trained meta learner.

Please note that to evaluate the performance scores of the stacking ensemble model using nested CV, the described procedure is repeated multiple times, depending on the number of outer loops in nested CV.

## Appendix E

The nested CV procedure consists of an outer loop for model evaluation while an inner loop tunes the hyperparameters. The performance estimates are calculated by averaging the test scores obtained across the different dataset splits in the outer loop. Varma and Simon [2] demonstrated that this approach, which avoids pooling the training and test data, produces nearly unbiased performance estimates. To further reduce bias and obtain more accurate estimates, the nested CV procedure could be repeated multiple times using different random shuffles of the labeled data. In our analysis, we utilized three random shuffles of nested CV with 10 outer loops and 5 inner loops, ensuring a robust assessment of the models’ performance. We remark that in order to conduct the hyperparameter tuning procedure in the inner loop of the nested CV, we used the Optuna framework [17]. Optuna is a powerful optimization library that efficiently searches for the optimal hyperparameters of the models. For more details on the nested cross-validation procedure, including information on determining the number of outer and inner loops and the computational implications, please refer to the online article by Jason Brownlee [31].

## Appendix F

**Figure 9:**
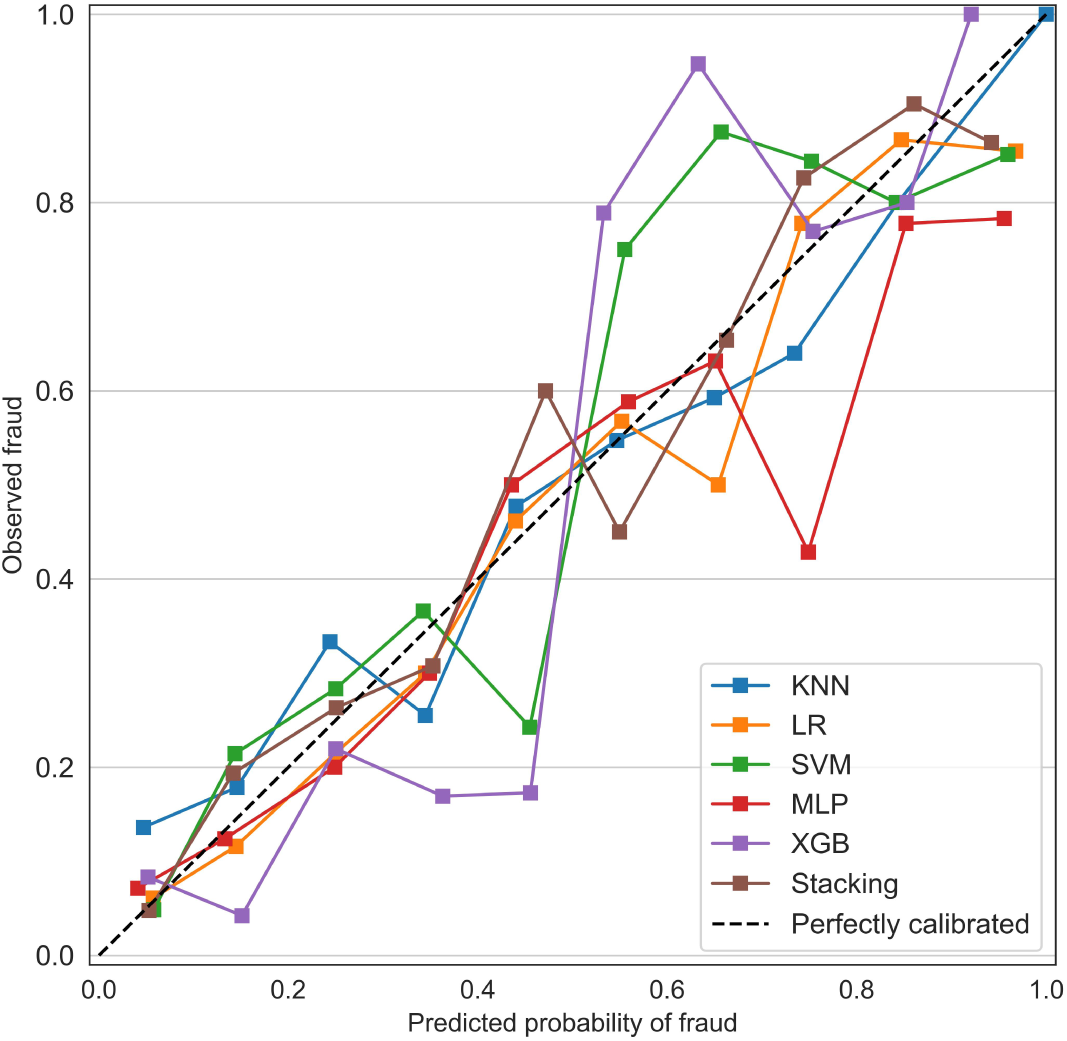
Calibration plots. KNN (blue line) indicates k-nearest neighbor; LR (orange line), logistic regression; SVM (greed line), support vector machines; MLP (red line), multilayer perceptron; XGB (purple line), extreme gradient boosting; and Stacking (brown line), stacking ensemble.

## Appendix G

### Overutilization Statistics for Medicare Jurisdictions

Below, we present the details of overutilization statistics computed for each Medicare jurisdiction. (Note that 0.0 as p-value means a value significantly smaller than 0.01.)

**Figure 10:**
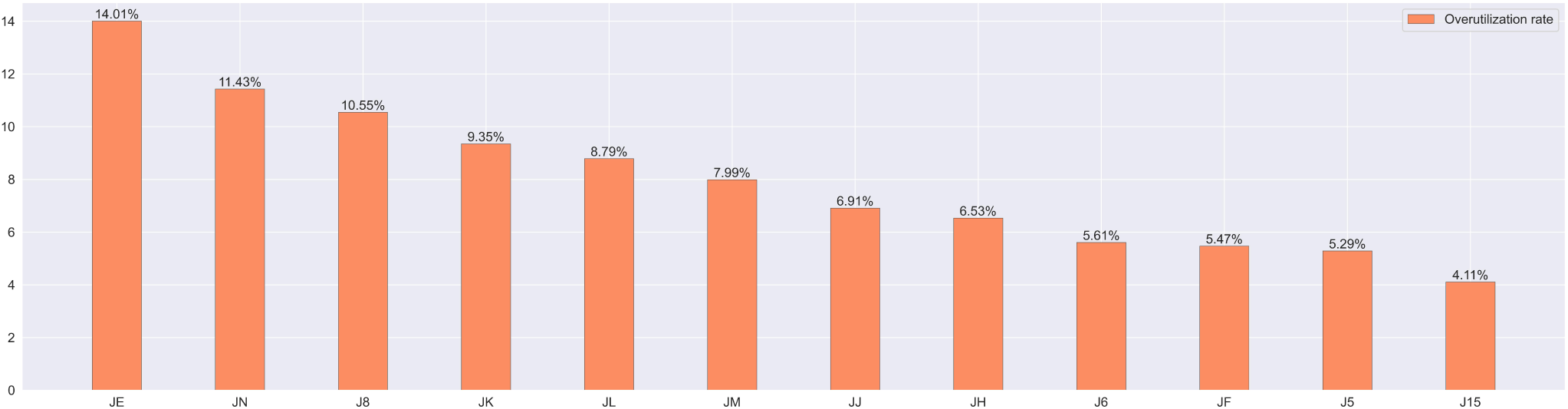
Predicted overutilization rate within Medicare jurisdictions in 2021.

**Table 7:** Overutilization rate within Medicare jurisdictions details.

|  | JE | JF | JH | JL | J5 | J8 | JJ | JM | JK | J6 | J15 | JN |
| --- | --- | --- | --- | --- | --- | --- | --- | --- | --- | --- | --- | --- |
| Overutilizer # | 301 | 68 | 140 | 163 | 35 | 87 | 64 | 101 | 234 | 70 | 32 | 162 |
| Non-fraud # | 1848 | 1175 | 2003 | 1692 | 627 | 738 | 862 | 1163 | 2268 | 1178 | 747 | 1255 |
| overutilization rate | 14.01% | 5.47% | 6.53% | 8.79% | 5.29% | 10.54% | 6.91% | 7.99% | 9.35% | 5.61% | 4.11% | 11.43% |

**Figure 11:**
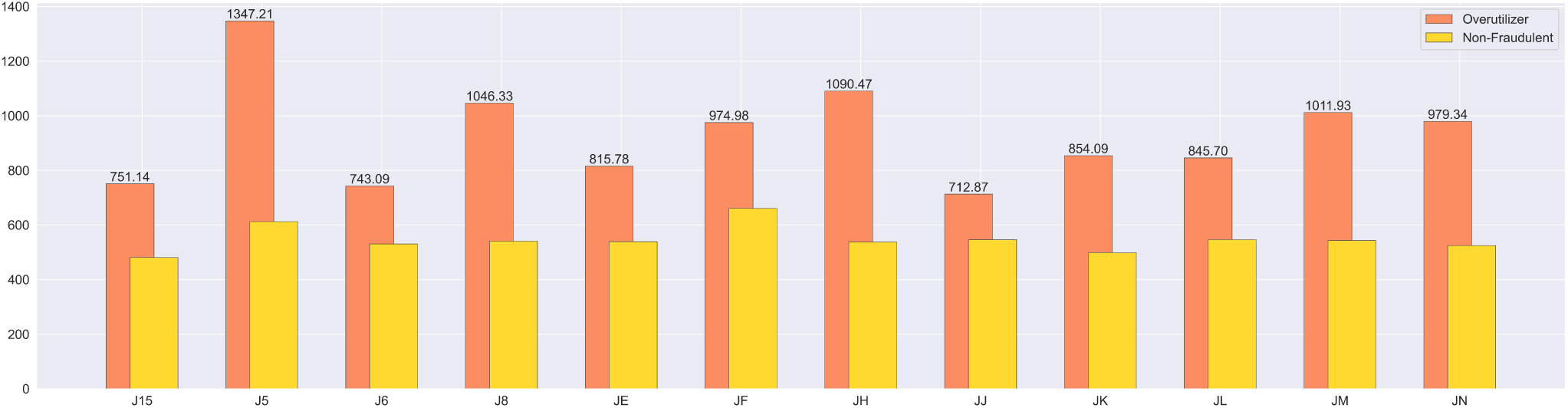
Comparing the average payment per patient between overutilizer and non-fraudulent ophthalmologists within Medicare Jurisdictions 2021.

**Table 8:**
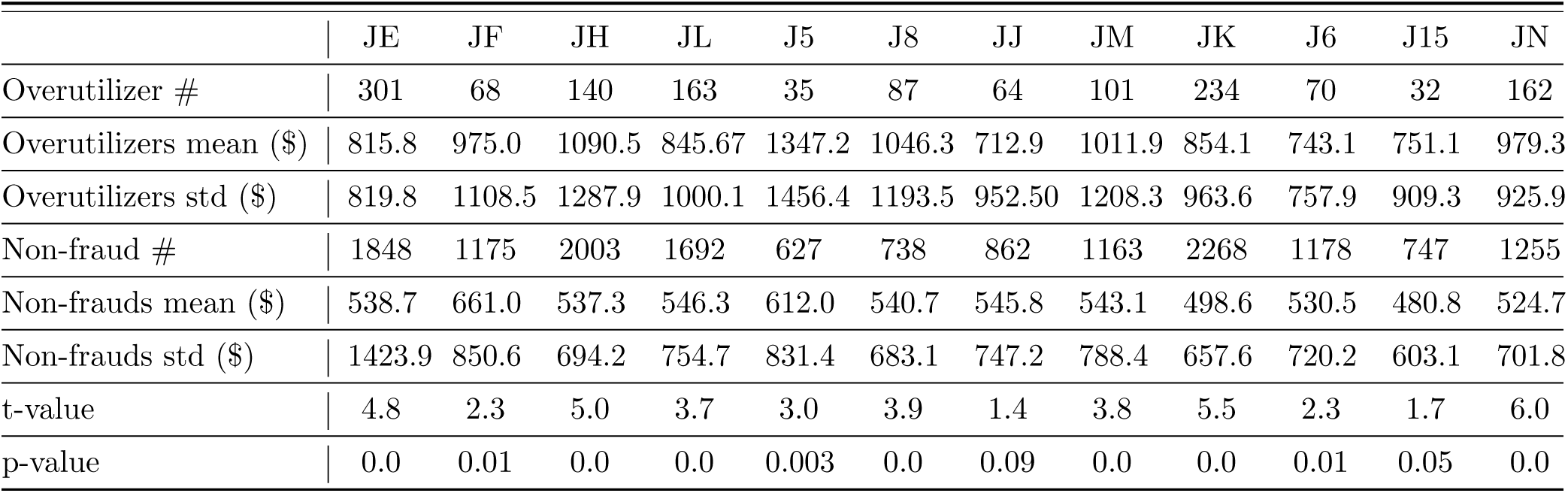
Average payment per patient within Medicare jurisdictions details.

**Figure 12:**
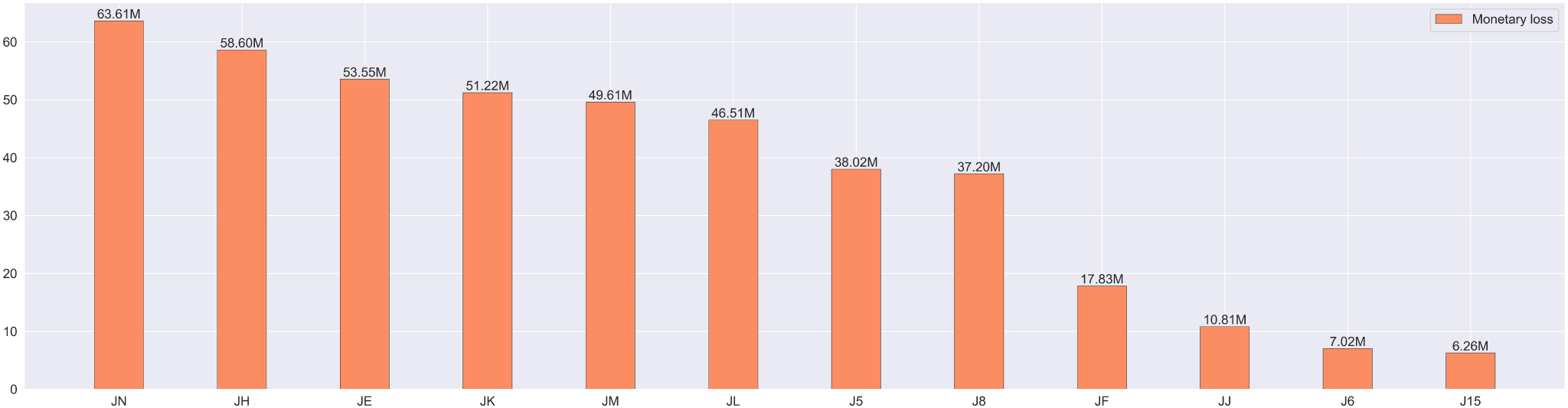
Monetary loss within Medicare jurisdictions in 2021.

**Table 9:** Monetary loss within Medicare jurisdictions details.

|  | JE | JF | JH | JL | J5 | J8 | JJ | JM | JK | J6 | J15 | JN |
| --- | --- | --- | --- | --- | --- | --- | --- | --- | --- | --- | --- | --- |
| Overutilizers total payment (\$m) | 150.5 | 43.5 | 104.0 | 108.6 | 54.2 | 66.1 | 35.8 | 91.8 | 117.0 | 27.0 | 13.6 | 130.3 |
| Monetary loss (\$m) | 53.5 | 17.8 | 58.6 | 46.5 | 38.0 | 37.2 | 10.8 | 49.6 | 51.2 | 7.0 | 6.2 | 63.6 |

### Non-Drug Medicare Payment Per Patient

The significant variation in Medicare payment per patient, as indicated by the high standard deviation calculated earlier, is primarily attributed to the inclusion of drug payments. If we exclude drug payments from the total payments, we anticipate that the average payment per patient for non-drug services among overutilizing providers will still be considerably higher than that of non-fraudulent providers. Below, we detail this statistic across the nation and within Medicare jurisdictions (See Figure 13 and Table 10.)

**Figure 13:**
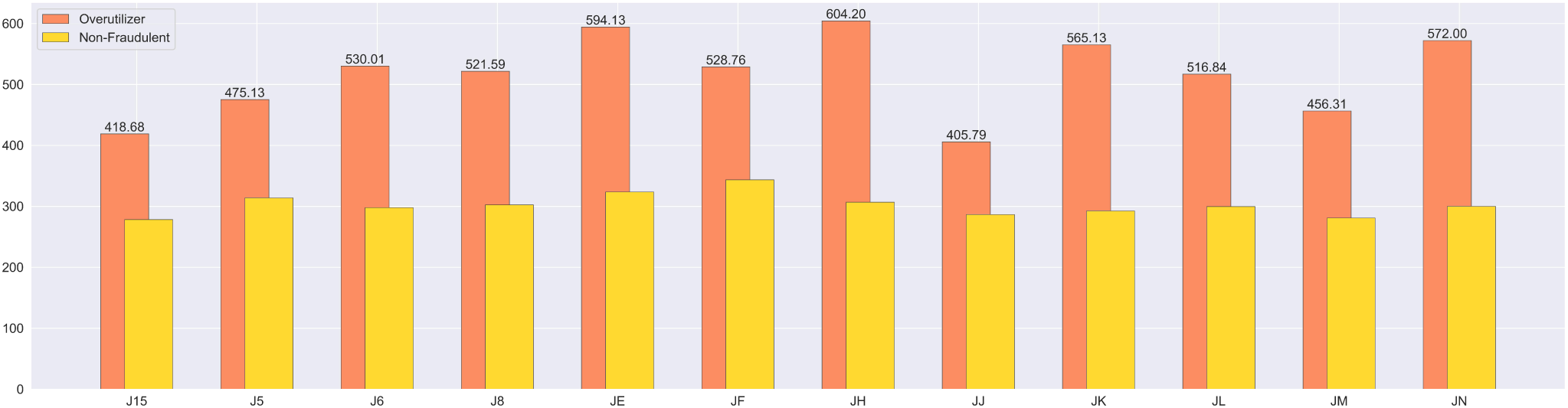
Comparing the average non-drug Medicare payment per patient between overutilizer and non-fraudulent ophthalmologists within Medicare Jurisdictions 2021.

**Table 10:** Average non-drug Medicare payment per patient within Medicare jurisdictions details.

|  | JE | JF | JH | JL | J5 | J8 | JJ | JM | JK | J6 | J15 | JN |
| --- | --- | --- | --- | --- | --- | --- | --- | --- | --- | --- | --- | --- |
| Overutilizer # | 301 | 68 | 140 | 163 | 35 | 87 | 64 | 101 | 234 | 70 | 32 | 162 |
| Overutilizers mean (\$) | 594.1 | 528.8 | 604.2 | 516.8 | 475.1 | 521.6 | 405.8 | 456.3 | 565.1 | 530.0 | 418.7 | 572.0 |
| Overutilizers std (\$) | 349.6 | 219.0 | 766.6 | 235.9 | 220.0 | 203.4 | 171.4 | 178.1 | 404.8 | 370.7 | 191.0 | 385.7 |
| Non-fraud # | 1848 | 1175 | 2003 | 1692 | 627 | 738 | 862 | 1163 | 2268 | 1178 | 747 | 1255 |
| Non-frauds mean (\$) | 323.4 | 343.4 | 306.7 | 299.5 | 313.8 | 302.4 | 286.6 | 280.7 | 292.4 | 297.9 | 278.3 | 300.0 |
| Non-frauds std (\$) | 174.1 | 175.9 | 157.9 | 149.6 | 149.5 | 150.3 | 133.5 | 140.4 | 148.3 | 174.4 | 134.6 | 159.5 |
| t-value | 11.9 | 6.2 | 4.2 | 10.4 | 4.0 | 8.7 | 4.7 | 8.2 | 9.3 | 5.0 | 3.8 | 7.9 |
| p-value | 0.0 | 0.0 | 0.0 | 0.0 | 0.0 | 0.0 | 0.0 | 0.0 | 0.0 | 0.0 | 0.0 | 0.0 |

Across the nation, the 1,457 overutilizing/fraudulent ophthalmologists (mean=$545.4, std=$388.9) compared to 15,556 non-fraudulent ophthalmologists (mean=$302.9, std=$157.1) had significantly higher non-drug Medicare payment per patient (t(17011)=21.3, p << 0.01).

## Appendix H

To compute performance metrics such as accuracy, specificity, precision, and sensitivity (recall), a specific probability threshold is required. Typically, these metrics are computed using a default threshold of 0.5, but this may not be optimal for imbalanced data situations.[3] Additionally, depending on the application, the emphasis may be placed on either precision or sensitivity, which would warrant a higher or lower threshold, respectively.

In the case of Medicare overutilization/fraud, if the primary goal is to avoid missing instances of overutilization/fraud, a higher sensitivity is desired, leading to a lower probability threshold. This approach would detect more providers as overutilizer/fraudulent, but it carries the risk of mislabeling non-fraudulent providers, potentially damaging their reputation if publicly announced. However, if the goal is to minimize mislabeling, a higher precision is preferred, requiring a higher probability threshold. While this is beneficial for public disclosure of overutilizing/fraudulent providers, it poses a financial burden on Medicare, as more instances of abuse go undetected.

In this study, we aim to strike a balance between precision and sensitivity. Thus, we seek a probability threshold that achieves a trade-off between these metrics. The F-score is optimized to determine such a balance, and the threshold is adjusted accordingly to maximize the average F-score obtained through the repeated nested CV procedure. This results in a probability threshold of 0.353. Accordingly, providers with a predicted overutilization/fraud probability exceeding 35.3% are labeled as overutilizer/fraudulent, while those below this threshold are considered non-fraudulent. The accuracy, specificity, precision, and sensitivity of the stacking ensemble model obtained using the probability threshold of 0.353 are presented in Table 11. The metrics are calculated via the repeated nested CV procedure.

**Table 11:** Additional performance metrics for the stacking ensemble model utilizing a probability threshold of 0.353.

| Accuracy (95% CI) | Specificity (95% CI) | Precision (95% CI) | Sensitivity (95% CI) |
| --- | --- | --- | --- |
| 0.850 (0.836-0.862) | 0.886 (0.867-0.903) | 0.750 (0.720-0.779) | 0.763 (0.733-0.793) |

### Reducing Mislabeling

Modifying the probability threshold not only would change the performance metrics, but also would lead to alterations in predicted overutilization statistics. In what follows, we adjust the probability threshold so as to reduce mislabeling. Subsequently, we recalculate the overutilization statistics and compare them with the results mentioned in the article.

To reduce mislabeling, we adjust the probability threshold to a higher value, resulting in an increased specificity (or a decreased false positive rate). Suppose our target is to achieve a specificity score of 95% or higher (or a false positive rate of 5% or lower). According to Figure 14, we can identify a probability threshold that maintains the same accuracy level while achieving a very low false positive rate. Specifically, we choose the probability threshold of 0.646. The corresponding performance metrics are presented in Table 12. The results demonstrate that the new probability threshold leads to higher specificity and precision, albeit at the expense of lower sensitivity. This choice of probability threshold is particularly suitable in situations where the main objective is to minimize false positives.

**Figure 14:**
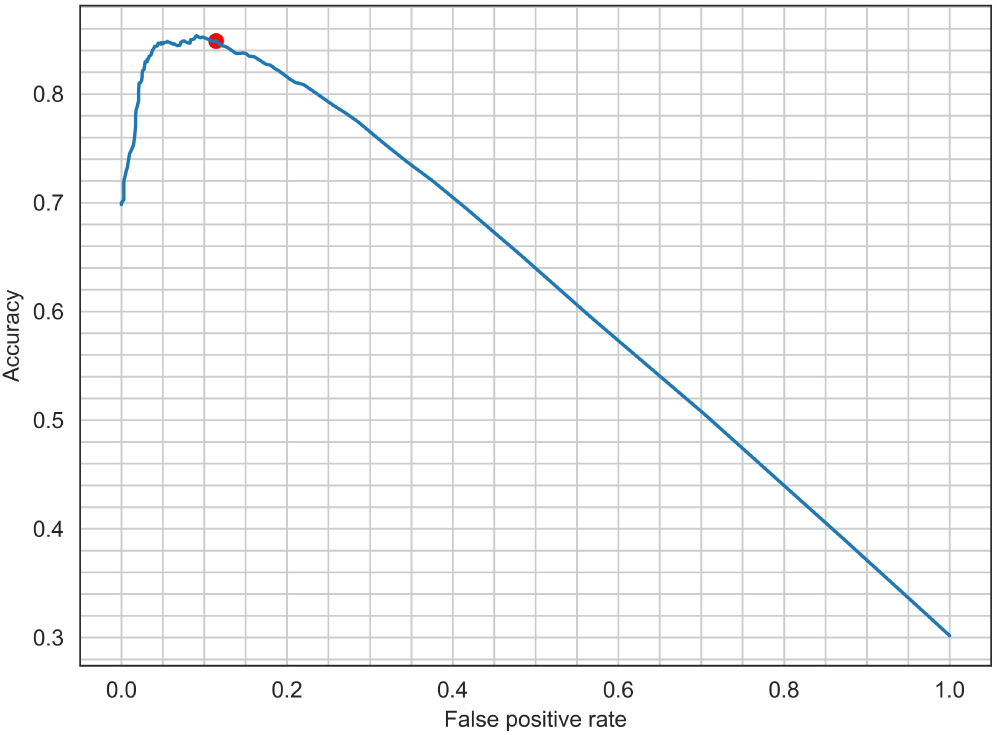
The Relation between the accuracy and false positive rate. The red dot corresponds to the probability threshold of 0.353.

**Table 12:** Performance metrics for the stacking ensemble model utilizing a probability threshold of 0.646.

| Accuracy (95% CI) | Specificity (95% CI) | Precision (95% CI) | Sensitivity (95% CI) |
| --- | --- | --- | --- |
| 0.847 (0.835-0.860) | 0.952 (0.939-0.965) | 0.854 (0.819-0.889) | 0.605 (0.5685-0.642) |

With this higher probability threshold, we expect to detect less overutilizer physicians, and therefore the estimated overutilization rate should decrease. Indeed, the estimated nationwide overutilization rate is decreased to 4.2% (from 8.6%), which still falls between the 3-10% fraud rate estimation [5], and the estimated monetary loss is decreased to $168.5 million (from $437.1 million). Additionally, Figure 15 shows the updated heat map of overutilization rate across the US.

**Figure 15:**
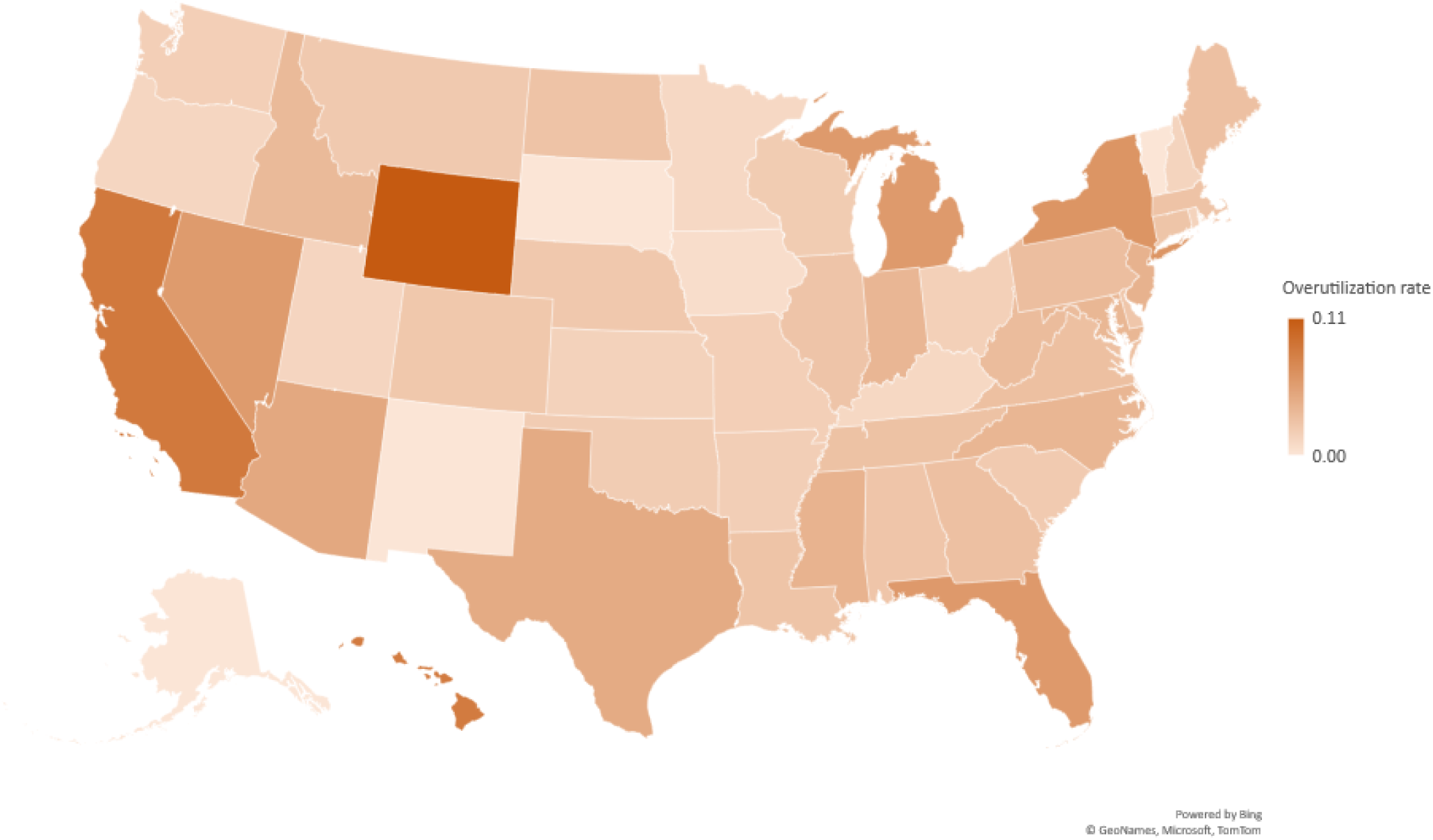
The heat map showcases the predicted overutilization rates across states in the United States. The rates are estimated using the probability threshold of 0.646.

It is intriguing to find that a state like CA, which previously had the highest overutilization rate (14.2%), does not have the highest rate in the new estimations. Instead, its overutilization rate is estimated to be 8.6% whereas the highest rate is 11.1%. The shift in the estimated overutilization rates for CA and many other states indicates that many physicians in these states fall into the category of borderline physicians. This means that the probability of them being overutilizers lies above 35% (previous probability threshold) but below 64% (the new probability threshold). The presence of a significant number of borderline physicians in these states highlights the need for further investigation into their billing practices.

## Appendix I

### Similarity between overutilization and normal utilization

We compare the billing patterns of the labeled providers across multiple years with the aim of identifying any discernible differences between the billing patterns of overutilizer and non-fraudulent cases. We employ the signature transform technique [9]. The signature transform, also known as the path signature, is a mathematical tool that processes sequential data such as time series or paths. It represents data points as higher-order tensors, capturing essential information about the data and associated features in a concise and expressive form.

For each provider, we extracted four aggregate features per year from 2013 to 2019: total services, total beneficiaries, total Medicare payment amount, and total Medicare drug payment amount. We computed the signature transform of the extracted time series data for each provider using the Signatory v1.2.6.1.9.0 package [23], resulting in tensors of length 84 for each provider. In order to visualize the high-dimensional data, we employ Principal Component Analysis (PCA) to reduce its dimensionality to 2. Figure 16 illustrates the plot of the first two principal components for overutilizer and non-fraudulent providers. The similarity between the overutilizer group and the non-fraudulent group is apparent, indicating the potential complexity and sophistication involved in fraudulent practices.

**Figure 16:**
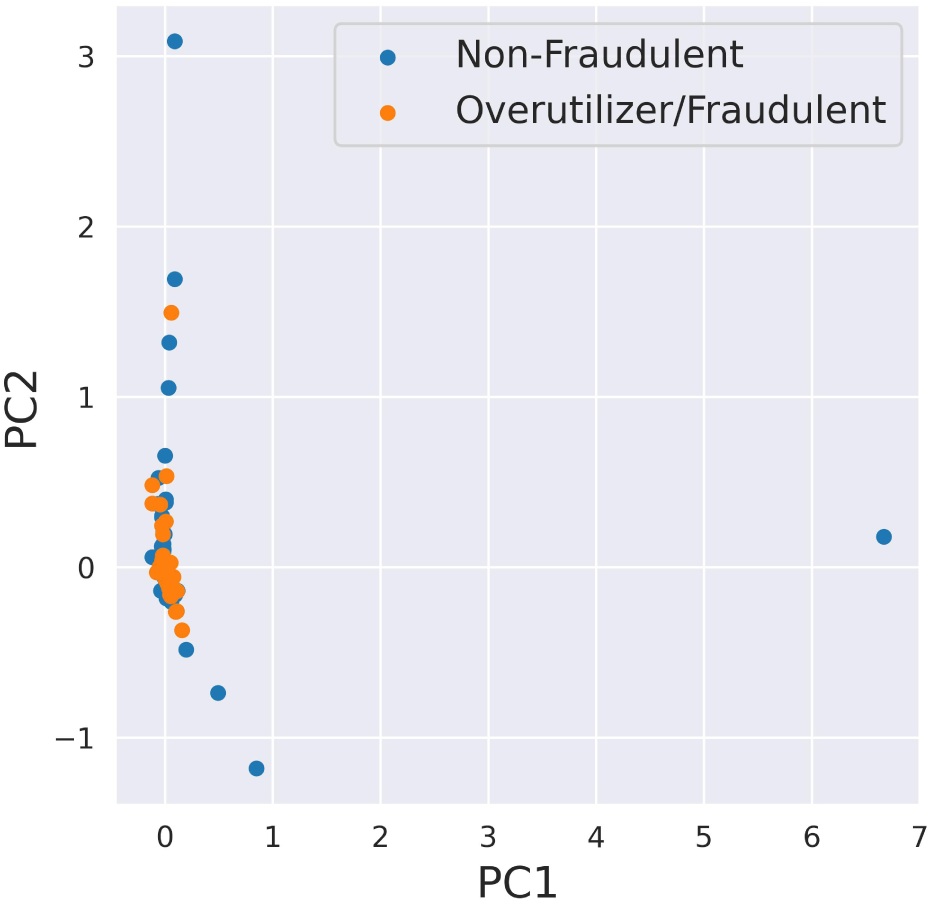
The PCA plot for the signature transforms. Each point on the plot represents a provider, with its position determined by the values of the first principal component and the second principal component obtained through the PCA analysis.

## Notes

### Competing Interest Statement

The authors have declared no competing interest.

### Funding Statement

This study did not receive any funding

### Author Declarations

1. https://data.cms.gov/provider-summary-by-type-of-service/medicare-inpatient-hospitals/medicare-inpatient-hospitals-by-provider-and-service 2. https://data.cms.gov/provider-summary-by-type-of-service/medicare-physician-other-practitioners/medicare-physician-other-practitioners-by-provider

